# Investigating the quality, trustworthiness and integrity of published randomized trials on phosphodiesterase type 5 inhibitors for the treatment of fetal growth restriction and their impact on study findings: protocol for a systematic review with aggregate data meta-analysis and data integrity assessments

**DOI:** 10.1101/2025.10.26.25338641

**Authors:** M.E. Bruins, E.M. Bordewijk, A. Pels, A.T. Papageorghiou, S.J. Gordijn, J.C. Jakobsen, J. Wilkinson, B.W. Mol, K.M. Groom, W. Ganzevoort

## Abstract

**Background:** Phosphodiesterase type 5 (PDE-5) inhibitors have been proposed as a potential in-utero treatment to improve uteroplacental perfusion in pregnancies with fetal growth restriction. Randomized controlled trials (RCTs) have shown conflicting results, with concerns about the integrity of several trials.

**Objective:** This protocol outlines a systematic review and aggregate data meta-analysis of RCTs investigating the short-term effects of PDE-5 inhibitors versus placebo or no treatment in pregnancies affected by FGR. We will assess the trustworthiness and scientific integrity of included trials and evaluate their impact on pooled outcomes.

**Methods:** Eligible trials were identified through a literature search. Primary outcome is mortality (stillbirth, neonatal or infant death). Secondary outcomes include severe neonatal morbidities and maternal preeclampsia. Risk of bias will be assessed using the Cochrane Risk of Bias 2 tool. Trustworthiness will be evaluated using the INSPECT-SR tool, the Cochrane Pregnancy and Childbirth Trustworthiness Screening Tool and additional statistical analyses. In the aggregate data meta-analysis, trials will be classified into subgroups based on the results from the INSPECT-SR tool. If concerns about trial conduct or reporting arise, authors will be contacted; unresolved issues may be referred to the journal.

**Discussion:** This review will critically assess the trustworthiness of randomized trials on PDE-5 inhibitors for FGR and summarize the available evidence.

This protocol is written in accordance with the Preferred Reporting Items for Systematic Review and Meta-Analysis Protocols (PRISMA-P) guidelines^1^. Corresponding PRISMA-P items are indicated in brackets following each section heading.

## INTRODUCTION

### Rationale {6}

Fetal growth restriction (FGR) affects 5-10% of pregnancies^3,4^ and is associated with adverse perinatal and long-term outcomes for the offspring.^5,6^ The most common underlying pathophysiological mechanism is uteroplacental insufficiency. There are currently no effective treatments to ameliorate the adverse *in utero* environment associated with uteroplacental insufficiency and FGR. Phosphodiesterase type 5 (PDE-5) inhibitors were proposed as the first-ever *in-utero* treatment for FGR, using the nitric oxide pathway affecting vascular resistance to improve uteroplacental perfusion. This hypothesis was supported by animal studies^7-11^, human *ex vivo*^12^ and small pre-clinical and clinical studies.^13-15^

Over the past decade, multiple randomized controlled trials have been conducted to investigate the therapeutic potential of PDE-5 inhibitors for the treatment of FGR, yielding conflicting findings. Some meta-analyses have suggested improvements in birth weight and pregnancy prolongation.^16,17^ However, the STRIDER trials^18-21^, considered high-quality studies, did not demonstrate any beneficial effects and described potential of harm,^20,22^ We are currently further investigating these findings through a systematic review and a pre-planned individual participant data meta-analysis, which will incorporate data from all four STRIDER trials (publication in preparation). Strikingly, investigators from 14 other eligible trials did not respond to requests to contribute to the individual participant data meta-analysis. The lack of inclusion of these other trials is likely to explain the conflicting results seen as the investigation of PDE-5 inhibitors as a potential *in-utero* treatment for FGR evolves. The unwillingness to contribute data, alongside retraction and expression of concerns for some published trials raises concern regarding the quality and scientific integrity of some of the trials in this field.^23,24^

Identifying studies at high risk of compromised integrity is crucial, as their inclusion in meta-analyses may influence the results, lead to unreliable conclusions, and potentially impact on clinical care that may be harmful. Given that some meta-analyses did not consider studies by their quality, trustworthiness and integrity, there is a need to evaluate the impact of including or excluding such studies. This approach will help to generate reliable and clinically meaningful evidence.

### Objectives {7}

The objective of this review is to conduct an aggregate data meta-analysis of randomised controlled trials on exploring the short-term effects of PDE-5 inhibitors, in comparison to placebo or no treatment, for the treatment of FGR. We will include a detailed analysis on trustworthiness of individual trials and evaluate the aggregate effects of based on trustworthiness.

## METHODS

This study will perform an aggregate data meta-analysis of randomized controlled trials identified through a previous systematic review, alongside a comprehensive assessment of the trustworthiness of each included trial.

The trials to be included have been identified through an earlier systematic review conducted as part of an individual participant data meta-analysis.^2^ For clarity, the eligibility criteria, information sources and selection process are outlined below.

### Eligibility criteria {8}

- Study type: randomized clinical trials
- Participants: women with singleton pregnancies affected by FGR (as defined by individual trials)
- Intervention: any PDE-5 inhibitor at any dose and by any route of administration with the intention of multiple dose administration
- Control: placebo or no intervention

### Information sources {9}

#### Databases

OVID MEDLINE, OVID EMBASE, the Cochrane Controlled Register of Trials (CENTRAL), and the clinical trial registers Clinicaltrials.gov and World Health Organization International Clinical Trials Registry Platform (ICTRP) from 1946 to present.

#### Grey literature

We included conference abstracts published in these databases. We did not hand search conference proceedings. We cross-checked the reference lists and the cited articles of relevant papers for additional relevant trials.^2^ We also included references cited in systematic review reports on the same or similar topic and from online searches on the topic.

#### Date of searches

Literature searches on the databases were conducted on 18 November 2019, with subsequent updates on 17 September 2020 and 25 September 2024. Grey literature was searched until March 2025.

### Search strategy {10}

The full search strategy is provided in Supplementary Appendix B.

### Selection process {11b}

Paired independent reviewers screened all titles and abstracts, advancing any citation deemed potentially eligible by either reviewer to full-text review. Full-text reviews were conducted independently by different paired reviewers. Discrepancies were resolved through discussion and, if necessary, by involving a third investigator.

### Data management {11a}

Records retrieved from database searches were managed in EndNote, where duplicates were removed. Title and abstract screening was performed using Rayyan or Microsoft Excel. Full-text was documented using standardized forms in Excel. Quality assessments and extracted data will be documented in the same way.

### Data collection process {11c}

Data on trial characteristics and outcomes will be extracted from the published articles. For trials in which individual participant data are available but outcomes were not reported in the publication, aggregate-level data will be derived from the individual participant data using SPSS software. Data extraction will be conducted using an artificial intelligence-based data extraction tool, which will import the information into a standardized Excel form. All extracted data will then be manually verified by a single human reviewer to ensure accuracy.

Data integrity assessments will be conducted in parallel with data extraction, following a predefined structured plan (below). Results will be documented in Excel.

### Data items {12}

#### Trial characteristics

- First author
- Title of the study
- Year of publication
- Inclusion criteria (including diagnosis of FGR and gestational age at inclusion)
- Exclusion criteria
- Control
- Intervention
- Third arm (if applicable)
- Co-intervention
- Intervention – sample size
- Control – sample size

#### Outcomes among all pregnancies

- Gestational age at birth
- Birth weight (grams)
- Maternal preeclampsia (as defined by individual trials)
- Stillbirth

#### Outcomes among liveborn

- Intraventricular haemorrhage (Papile grade 3 or 4, or as defined by individual trials)
- Cystic periventricular leukomalacia (grade two or more, or as defined by individual trials)
- Bronchopulmonary dysplasia (as defined by individual trials)
- Necrotising enterocolitis requiring surgery (as defined by individual trials)
- Retinopathy of prematurity requiring treatment (as defined by individual trials)
- Neonatal and infant death

#### For data integrity assessments

The following checks and methodological references for the data integrity assessments are based on the scoping review by *Bordewijk et al*. and the INSPECT-SR (INveStigating ProblEmatic Clinical Trials in Systematic Reviews) tool.^25^ (available via: https://osf.io/b74wj).

Trial characteristics

- Trial registry
- Data availability and transparency (based on the willingness of authors to share individual participant data)
- Single or multicenter trial
- Number of participants included
- Number of participants lost to follow up
- Timespan of recruitment
- Journal of publication of the results
- Date received by journal
- Date start recruitment
- Date trial registry

Author characteristics

- Retractions or expressions of concern involving any of the authors of the included paper (if so, the reason) using Retraction Watch Database^26^
- Number of published randomized controlled trials authored or co-authored by the first author of the included paper
- Search with Google Scholar for similar articles or abstracts by the same author group

Statistical methods

Statistical analyses will be conducted independently by two reviewers.

- Plausibility and correctness of reported statistics
  - Reproducibility of the P-values as reported by the authors
  - Reproducibility of the effect sizes and confidence intervals
  - Distribution of the P-values of the baseline characteristics, split for continuous and dichotomous tests

Other

- Internal consistency
- Registry concordance
- Chronological plausibility
- Plagiarism check
- Duplication of numbers with other publications
- Image duplication
- Feasibility and plausibility (governance, methodology, execution, results, reporting)

### Outcomes and prioritization {13}

#### Primary outcome^2^

- Mortality (defined as either stillbirth, neonatal or infant mortality)

#### Secondary outcomes

- Cerebral intraventricular haemorrhage (Papile grade 3 or 4^27^, or as defined by individual trials)
- Cystic periventricular leukomalacia (grade two or more^28^, or as defined by individual trials)
- Bronchopulmonary dysplasia (as defined by individual trials)
- Necrotising enterocolitis requiring surgery (as defined by individual trials)
- Retinopathy of prematurity requiring treatment (as defined by individual trials)
- Maternal preeclampsia

Exploratory outcomes

- Gestational age at birth
- Birth weight (grams)
- Stillbirth
- Neonatal and infant death (among liveborn infants, as defined as by individual trials)

### Quality assessment

Multiple types of quality assessments will be conducted for all eligible trials.

#### Risk of bias {14}

Risk of bias will be assessed using version 2 of the Cochrane tool for assessing risk of bias in randomised trials (RoB 2)^29^. The tool assesses five domains 1) bias arising from the randomization process 2) bias due to deviations from intended interventions 3) bias due to missing outcome data 4) bias in measurement of the outcome and 5) bias in selection of the reported result. Individual trials will be classified as:

- Low risk
- Some concerns
- High risk

#### Trustworthiness

Trustworthiness and data integrity will be checked using the INSPECT-SR (INveStigating ProblEmatic Clinical Trials in Systematic Reviews) tool^30^ (available via: https://osf.io/b74wj). INSPECT-SR includes 21 checks in four domains 1) inspecting post-publication notices (3 checks), 2) inspecting conduct, governance, and transparency (5 checks), 3) inspecting text and figures (2 checks), 4) inspecting results in the study (11 checks). Individual trials will be classified as:

- No concerns
- Some concerns
- Serious concerns

In addition, it will be assessed using the Trustworthiness Screening Tool developed by the Cochrane Pregnancy and Childbirth Group^31^. The tool assesses four domains 1) research governance 2) plausibility of baseline characteristics 3) feasibility and 4) plausibility of results. Individual trials will be classified as:

- Excluded
- Awaiting classification
- Included

Lastly, statistics will be checked using an existing spreadsheet that recalculates P-values based on reported means, standard deviations (t-tests) and absolute numbers (chi-square tests). Baseline characteristics will also be assessed for statistically significant differences.

#### Notifying the authors

If indicators of integrity issues are identified, the primary contact of the trial will be notified by email, requesting clarification of unclear aspects of the methodology or results. If an inadequate or no response is received, a letter requesting an investigation and, depending on the result of the investigation, retraction will be sent to the journal in which the article was published. Examples of both letters are included in the Supplementary Appendix C.

#### Subgroups based on quality assessments

Based on the results of the INSPECT-SR, subgroups (no concerns, some concerns or serious concerns) will be defined for the aggregate data meta-analysis.

### Data synthesis {15}

#### Criteria under which trial data will be quantitatively synthesised {15a}

All available data will be included in the meta-analysis, which will be conducted without the application of predefined eligibility thresholds.

#### Statistical approach {15b}

In accordance with our original protocol^2^, the aggregate data meta-analyses will be conducted according to the Cochrane Handbook of Systematic Reviews of Interventions^32^, Keus et al.^33^, and the eight-step assessment proposed by Jakobsen et al^34^. Subgroup analyses will be performed based on the results of the trustworthiness and integrity assessments (see Quality assessment). Intervention effects will be assessed using both random-effects and fixed-effect meta-analyses for each treatment comparison, with the analysis with the highest P-value being used as main result.^32-34^ A P-value of 0.02 or less will be considered statistically significant.^34^ Heterogeneity among trials will be identified by visual inspection of forest plots and by calculating the *I*^*2*^ value.^32^

Data from intention-to-treat population will be used where possible. However, for participants with missing outcome data, analysis will be based on the available data, without imputing missing values. In a multiple-arm trial, if two or more intervention groups are relevant and included as separate comparisons in the same meta-analysis, and they share a common control group, the control group should be split (e.g. halved) to avoid double counting participants. However, if only one intervention group is relevant to the review or meta-analysis, the study can be treated as a standard two-arm trial, and the full control group can be used without adjustment^32^. Descriptive statistics (mean, standard deviation (SD), numbers and proportions) will be used to describe the patient and intervention related characteristics. Binary outcomes will be reported as risk ratios and absolute risk differences with 95% confidence intervals, while continuous outcomes will be reported as mean differences with standard deviations or as medians with interquartile ranges. The statistical analysis will be performed using the current version of R.

#### Additional analyses {15c}

The impact of missing data will be assessed in prespecified sensitivity analyses^2^ using the ‘best-worst-case’ scenario and the ‘worst-best-case’ scenario.^34^ In case of continuous outcomes, standard deviations will be imputed from P-values according to the Cochrane Handbook for Systematic Reviews of Intervention.^30^ If it was not possible to calculate the standard deviation from the P value or the confidence intervals, the highest standard deviation from the other trials that included the relevant outcome will be imputed.

Clinical interpretations will be limited to trials meeting trustworthiness criteria. Meta-analyses including trials that do not meet these criteria will be conducted exclusively to explore their potential effect on the overall results.

### Meta-bias {16}

Where more than 10 trials are included, we will visually inspect funnel plots to assess for reporting bias.^2^

### Confidence in cumulative evidence {17}

Confidence in cumulative evidence will not be formally assessed using GRADE, as this review does not focus on evaluating intervention effects or clinical outcomes, but instead aims to investigate the impact of data integrity on subgroup analyses.

## ADMINISTRATIVE INFORMATION

### Registration {2}

PROSPERO (CRD42017069688)

### Author contributions {3b}

All authors contributed to the development of the protocol. WG conceptualized the review question. WG and MB designed the methodology, which was refined with input from all authors. WG and KMG were involved in the design of the search strategy. EMB designed the data integrity assessments. MEB drafted the initial version of the protocol with contributions from all authors.

### Amendments {4}

The search strategy was conducted in advance in accordance with a previously published protocol, that included individual participant data meta-analysis, as well as aggregate data meta-analysis^2^. Authors of eligible trials were approached to contribute to the individual participant data meta-analysis. However, the majority of authors did not respond to the invitation to share data. This prompted a study plan for a structured analysis of the trustworthiness of all eligible published trials. The current study will report on the aggregate data meta-analysis, with some minor amendments that have been incorporated into the current version of the protocol and are described in Supplementary Appendix A. Due to limited data availability, the prespecified outcomes have been revised to improve the feasibility of the study and ensure the potential for meaningful analysis. Additionally, a further investigation will be conducted to assess the trustworthiness and scientific integrity of the included trials. The results of this assessment will be incorporated as subgroups in the aggregate data meta-analysis.

### Support {5}

This study will be conducted without financial support.

## DECLARATIONS

### Author approval

All authors have seen and approved the manuscript.

### Data availability statement

Not applicable as no new data is generated.

## References

1. Moher D, Shamseer L, Clarke M, et al. Preferred reporting items for systematic review and meta-analysis protocols (PRISMA-P) 2015 statement. Syst Rev 2015;4(1):1. DOI: 10.1186/2046-4053-4-1.

2. Liauw J, Groom K, Ganzevoort W, et al. Short-term outcomes of phosphodiesterase type 5 inhibitors for fetal growth restriction: a study protocol for a systematic review with individual participant data meta-analysis, aggregate meta-analysis, and trial sequential analysis. Syst Rev 2021;10(1):305. DOI: 10.1186/s13643-021-01849-5.

3. Nardozza LMM, Caetano ACR, Zamarian ACP, et al. Fetal growth restriction: current knowledge. Arch Gynecol Obstet 2017;295(5):1061-1077. (In English). DOI: 10.1007/s00404-017-4341-9.

4. Lindqvist PG, Molin J. Does antenatal identification of small-for-gestational age fetuses significantly improve their outcome? Ultrasound Obst Gyn 2005;25(3):258–264. (In English). DOI: 10.1002/uog.1806.

5. Murray E, Fernandes M, Fazel M, Kennedy SH, Villar J, Stein A. Differential effect of intrauterine growth restriction on childhood neurodevelopment: a systematic review. Bjog-Int J Obstet Gy 2015;122(8):1062-1072. (In English). DOI: 10.1111/1471-0528.13435.

6. von Beckerath A-K KM, Rotky-Fast C, Karpf E, Lang U, Klaritsch P. Perinatal complications and long-term neurodevelopmental outcome of infants with intrauterine growth restriction. American Journal of Obstetrics and Gynecology 2013;208:130.e1–130.e6.

7. Zoma WD, Baker RS, Clark KE. Effects of combined use of sildenafil citrate (Viagra) and 17beta-estradiol on ovine coronary and uterine hemodynamics. Am J Obstet Gynecol 2004;190(5):1291–7. DOI: 10.1016/j.ajog.2003.12.021.

8. Stanley JL, Andersson IJ, Poudel R, et al. Sildenafil citrate rescues fetal growth in the catechol-O-methyl transferase knockout mouse model. Hypertension 2012;59(5):1021–8. DOI: 10.1161/HYPERTENSIONAHA.111.186270.

9. Satterfield MC, Bazer FW, Spencer TE, Wu G. Sildenafil citrate treatment enhances amino acid availability in the conceptus and fetal growth in an ovine model of intrauterine growth restriction. J Nutr 2010;140(2):251–8. DOI: 10.3945/jn.109.114678.

10. Oyston C, Stanley JL, Oliver MH, Bloomfield FH, Baker PN. Maternal Administration of Sildenafil Citrate Alters Fetal and Placental Growth and Fetal-Placental Vascular Resistance in the Growth-Restricted Ovine Fetus. Hypertension 2016;68(3):760–7. DOI: 10.1161/HYPERTENSIONAHA.116.07662.

11. Dilworth MR, Andersson I, Renshall LJ, et al. Sildenafil citrate increases fetal weight in a mouse model of fetal growth restriction with a normal vascular phenotype. PLoS One 2013;8(10):e77748. DOI: 10.1371/journal.pone.0077748.

12. Wareing M, Myers JE, O’Hara M, Baker PN. Sildenafil citrate (Viagra) enhances vasodilatation in fetal growth restriction. J Clin Endocrinol Metab 2005;90(5):2550–5. DOI: 10.1210/jc.2004-1831.

13. von Dadelszen P, Dwinnell S, Magee LA, et al. Sildenafil citrate therapy for severe early-onset intrauterine growth restriction. BJOG 2011;118(5):624–8. DOI: 10.1111/j.1471-0528.2010.02879.x.

14. Dastjerdi MV, Hosseini S, Bayani L. Sildenafil citrate and uteroplacental perfusion in fetal growth restriction. J Res Med Sci 2012;17(7):632–6. (https://www.ncbi.nlm.nih.gov/pubmed/23798922).

15. Paauw ND, Terstappen F, Ganzevoort W, Joles JA, Gremmels H, Lely AT. Sildenafil During Pregnancy: A Preclinical Meta-Analysis on Fetal Growth and Maternal Blood Pressure. Hypertension 2017;70(5):998–1006. DOI: 10.1161/HYPERTENSIONAHA.117.09690.

16. Rakhanova Y, Almawi WY, Aimagambetova G, Riethmacher D. The effects of sildenafil citrate on intrauterine growth restriction: a systematic review and meta-analysis. BMC Pregnancy Childbirth 2023;23(1):409. DOI: 10.1186/s12884-023-05747-7.

17. Liu Y, Un EM, Bai Y, et al. Safety and efficacy of phosphodiesterase-5 (PDE-5) inhibitors in fetal growth restriction: a systematic literature review and meta-analysis. J Pharm Pharm Sci 2024;27:13206. DOI: 10.3389/jpps.2024.13206.

18. Sharp A, Cornforth C, Jackson R, et al. Maternal sildenafil for severe fetal growth restriction (STRIDER): a multicentre, randomised, placebo-controlled, double-blind trial. Lancet Child Adolesc Health 2018;2(2):93–102. DOI: 10.1016/S2352-4642(17)30173-6.

19. Groom KM, McCowan LM, Mackay LK, et al. STRIDER NZAus: a multicentre randomised controlled trial of sildenafil therapy in early-onset fetal growth restriction. BJOG 2019;126(8):997–1006. DOI: 10.1111/1471-0528.15658.

20. Pels A, Derks J, Elvan-Taspinar A, et al. Maternal Sildenafil vs Placebo in Pregnant Women With Severe Early-Onset Fetal Growth Restriction: A Randomized Clinical Trial. JAMA Netw Open 2020;3(6):e205323. DOI: 10.1001/jamanetworkopen.2020.5323.

21. von Dadelszen P, Audibert F, Bujold E, et al. Halting the Canadian STRIDER randomised controlled trial of sildenafil for severe, early-onset fetal growth restriction: ethical, methodological, and pragmatic considerations. BMC Res Notes 2022;15(1):244. DOI: 10.1186/s13104-022-06107-y.

22. Pels A, Onland W, Berger RMF, et al. Neonatal pulmonary hypertension after severe early-onset fetal growth restriction: post hoc reflections on the Dutch STRIDER study. Eur J Pediatr 2022;181(4):1709–1718. DOI: 10.1007/s00431-021-04355-x.

23. Pels A, Ganzevoort W, Kenny LC, et al. Interventions affecting the nitric oxide pathway versus placebo or no therapy for fetal growth restriction in pregnancy. Cochrane Database Syst Rev 2023;7(7):CD014498. DOI: 10.1002/14651858.CD014498.

24. Turner JM, Russo F, Deprest J, Mol BW, Kumar S. Phosphodiesterase-5 inhibitors in pregnancy: Systematic review and meta-analysis of maternal and perinatal safety and clinical outcomes. BJOG 2022;129(11):1817–1831. DOI: 10.1111/1471-0528.17163.

25. Wilkinson JD, Heal C, Flemyng E, et al. INSPECT-SR: a tool for assessing trustworthiness of randomised controlled trials. medRxiv 2025 (10.1101/2025.09.03.25334905).

26. Integrity TCfS. The Retraction Watch Database. New York: The Center for Scientific Integrity; 2018.

27. Papile LA, Burstein J, Burstein R, Koffler H. Incidence and evolution of subependymal and intraventricular hemorrhage: a study of infants with birth weights less than 1,500 gm. J Pediatr 1978;92(4):529–34. DOI: 10.1016/s0022-3476(78)80282-0.

28. de Vries LS, Eken P, Dubowitz LM. The spectrum of leukomalacia using cranial ultrasound. Behav Brain Res 1992;49(1):1–6. DOI: 10.1016/s0166-4328(05)80189-5.

29. Sterne JAC, Savovic J, Page MJ, et al. RoB 2: a revised tool for assessing risk of bias in randomised trials. BMJ 2019;366:l4898. DOI: 10.1136/bmj.l4898.

30. Wilkinson J, Heal C, Antoniou GA, et al. Protocol for the development of a tool (INSPECT-SR) to identify problematic randomised controlled trials in systematic reviews of health interventions. BMJ Open 2024;14(3):e084164. DOI: 10.1136/bmjopen-2024-084164.

31. Alfirevic ZK. F.J.; Weeks, J.; Stewart, F.; Jones, L.; Hampson, L. Identifying and handling potentially untrustworthy trials – Trustworthiness Screening Tool (TST) developed by the Cochrane Pregnancy and Childbirth Group. Version 3.0. (file:///C:/Users/P090207/Downloads/PGC%20TST%20FINAL%20(1).pdf).

32. Higgins JT, J.; Chandler, J.; Cumpston, M.; Li, T.; Page, MJ.; Welch, VA. Cochrane Handbook for Systematic Reviews of Interventions version 6.5. Updated August 2024 ed. Cochrane 2024.

33. Keus F, Wetterslev J, Gluud C, van Laarhoven CJHM. Evidence at a glance: error matrix approach for overviewing available evidence. Bmc Medical Research Methodology 2010;10 (In English). DOI: Artn 90 10.1186/1471-2288-10-90.

34. Jakobsen JC WJ, Winkel P, Lange T, Gluud C. Thresholds for statistical and clinical significance in systematic reviews with meta-analytic methods. BMC Medical Research Methodology 2014;14:120. DOI: 10.1186/1471-2288-14-120.

35. Ayres-de-Campos D, Spong CY, Chandraharan E, Panel FIFMEC. FIGO consensus guidelines on intrapartum fetal monitoring: Cardiotocography. Int J Gynaecol Obstet 2015;131(1):13–24. DOI: 10.1016/j.ijgo.2015.06.020.

