## Supplementary Appendix A for "Investigating the quality, trustworthiness and integrity of published randomized trials on phosphodiesterase type 5 inhibitors for the treatment of fetal growth restriction and their impact on study findings: protocol for a systematic review with aggregate data meta-analysis and data integrity asses"

**Supplementary Appendix A – Protocol deviations**

The study protocol for a systematic review with individual participant data meta-analysis, aggregate meta-analysis, and trial sequential analysis was published[2] and registered in PROSPERO (#CRD42017069688, amended in October 2020).

Following the systematic review, we contacted the primary contacts of eligible trial registries and publications responded to the request for individual participant data. As no contacts responded (initially or after failing further email contact), the decision was taken to deviate from the protocol and present the results of the individual participant data meta-analysis in a separate publication. Additionally, we added the detailed data integrity assessments and subgroups analyses based on the results of the quality assessments.

**Additional deviations from the original protocol include the following:**

- The updated version of the *Cochrane Handbook for Systematic Reviews of Interventions* was used to guide methodological decisions.
- In addition to trials identified through our own literature search, we included trials referenced in other systematic reviews or encountered online that were not retrieved by our literature search strategy.
- Individual participant data were considered unattainable after two contact attempts, including one follow-up email sent one month after the initial request.
- Trial authors were not contacted to provide missing data for the aggregate data analysis, as they had not responded to the IPD request.
- Data extraction was conducted using an artificial intelligence tool, followed by verification by a single human reviewer, rather than by two independent human assessors as originally planned.
- Exploratory outcomes were added.
- Subgroup analyses were conducted based on trial quality assessments, rather than the originally specified subgroups.
- The GRADE (Grading of Recommendations, Assessment, Development and Evaluation) framework was not performed.
- A Summary of Findings table was not included.
