## Supplementary Appendix B for "Investigating the quality, trustworthiness and integrity of published randomized trials on phosphodiesterase type 5 inhibitors for the treatment of fetal growth restriction and their impact on study findings: protocol for a systematic review with aggregate data meta-analysis and data integrity asses"

**Supplementary Appendix B – Search strategy**

Ovid MEDLINE(R)
ALL <1946 to September 24, 2024>

| # | Search |
|---|---|
| 1 | fetal growth retardation/ |
| 2 | placental insufficiency/ |
| 3 | exp hypertension, pregnancy-induced/ |
| 4 | ((f?etal or f?etus* or intrauterin* or uterine or utero or antenat* or prenat* or early-onset or antenat* or ante-nat* or prenat* or pre-nat* or first trimester or 1st trimester) adj6 (growth adj3 (retard* or restrict*))).tw,kf. |
| 5 | (FGR* or IUGR*).tw,kf. |
| 6 | ((early or pregnancy or gestational) adj2 (growth retard* or growth restrict*)).tw,kf. |
| 7 | ((estimat* adj4 (f?etal or f?etus*) adj2 weight*) or EFW).tw,kf. |
| 8 | (((absent or revers*) adj6 (enddiastol or diastol* or doppler) adj2 (velocity or flow)) or AEDF* or A-EDF* or AREDF* or A-REDF*).tw,kf. |
| 9 | (abdom* adj3 circumfer*).tw,kf. |
| 10 | (placent* adj3 (insufficien* or d#sfunct* or disorder*)).tw,kf. |
| 11 | (((gestational or maternal or pregnancy) adj (hypertension or high blood pressur*)) or pregnancy-induced hypertension or (pregnancy adj3 hypertensive disorder*)).tw,kf. |
| 12 | (HELPP or preeclam* or eclam*).tw,kf. |
| 13 | or/1-12 [ FGR, placental insufficiency, pregnancy hypertensive disorders ] |
| 14 | phosphodiesterase 5 inhibitors/ or sildenafil citrate/ or tadalafil/ |
| 15 | (sildenafil* or revatio or homosildenafil or hydroxyhomosildenafil or Viagra or acetildenafil or desmethylsildenafil or NCX-911 or NCX911 or UK-92480* or UK92480* or tadalafil* or cialis or ic-351 or ic351).tw,kf. |
| 16 | (((((phosphodiesteras* or phospho-diesteras* or PDE) adj3 ("5" or V)) or phosphodiesterase5 or PDE5 or phosphodiesteraseV or PDEV) adj5 (inhib* or block* or antagonist* or anti)) or PD5i* or PD5-I or PDVi* or PDV-i).tw,kf. |
| 17 | or/14-16 [PD5-inhibitors] |
| 18 | 13 and 17 [ FGR & PD5-inhibitors ] |
| 19 | (TADAFER or (Strider not (short-tandem-repeat or DNA-strider* or ((water or oil or bike or bikes or biped*) adj5 strider*)))).ti. |
| 20 | 18 or 19 [ FGR & PD5-inhibitors -Strider/Tadafer trial ] |
| 21 | ((controlled clinical trial or randomized controlled trial).pt. or double-blind method/ or placebos/ or (random* or (controlled adj3 (study or trial)) or placebo* or double-blind*).tw,kf. or trial.ti.) not (("systematic review" or review).pt. or review.ti.) [ RCT-filter adapted from the Cochrane, reviews excluded ] |
| 22 | (exp animals/ not humans/) or ((pig or pigs or goat or goats* or sheep or lamb or lambs or ovine or rodent* or rabbit* or mice or mouse or murine* or rat or rats).ti. not human*.ti,ot.) [ animal filter] |
| 23 | 21 not 22 [ human RCT filter, adapted from the Cochrane ] |
| 24 | 20 and 23 [ human RCTs on FGR & PD5-inhibitors ] |
| 25 | remove duplicates from 24 [ human RCTs on FGR & PD5-inhibitors, deduplicated ] |

Embase
Source: Embase Classic+Embase <1947 to 2024 September 24>

| # | Search |
|---|---|
| 1 | intrauterine growth retardation/ |
| 2 | placenta insufficiency/ |
| 3 | maternal hypertension/ or exp "eclampsia and preeclampsia"/ |
| 4 | fetal doppler/ or abdominal circumference/ |
| 5 | ((f?etal or f?etus* or intrauterin* or uterine or utero or antenat* or prenat* or early-onset or antenat* or ante-nat* or prenat* or pre-nat* or first trimester* or 1st trimester*) adj6 (growth adj3 (retard* or restrict*))).tw,kw. |
| 6 | (FGR* or IUGR*).tw,kw. |
| 7 | ((early or pregnancy or gestational) adj2 (growth retard* or growth restrict*)).tw,kw. |
| 8 | ((estimat* adj4 (f?etal or f?etus*) adj2 weight*) or EFW).tw,kw. |
| 9 | (((absent or revers*) adj6 (enddiastol or diastol* or doppler) adj2 (velocity or flow)) or AEDF* or A-EDF* or AREDF* or A-REDF*).tw,kw. |
| 10 | (abdom* adj3 circumfer*).tw,kw. |
| 11 | (placent* adj3 (insufficien* or d#sfunct* or disorder*)).tw,kw. |
| 12 | (((gestational or maternal or pregnancy) adj (hypertension or high blood pressur*)) or pregnancy-induced hypertension or (pregnancy adj3 hypertensive disorder*)).tw,kw. |
| 13 | (HELPP or preeclam* or eclam*).tw,kw. |
| 14 | or/1-13 [ FGR, placental insufficiency, pregnancy hypertensive disorders ] |
| 15 | exp phosphodiesterase v inhibitor/ |
| 16 | (sildenafil* or revatio or homosildenafil or hydroxyhomosildenafil or Viagra or acetildenafil or desmethylsildenafil or NCX-911 or NCX911 or UK-92480* or UK92480* or tadalafil* or cialis or ic-351 or ic351).tw,kw. |
| 17 | (((((phosphodiesteras* or phospho-diesteras* or PDE) adj3 ("5" or V)) or phosphodiesterase5 or PDE5 or phosphodiesteraseV or PDEV) adj5 (inhib* or block* or antagonist* or anti)) or PD5i* or PD5-I or PDVi* or PDV-i).tw,kw. |
| 18 | or/15-17 [PD5-inhibitors] |
| 19 | 14 and 18 [ FGR & PD5-inhibitors ] |
| 20 | (TADAFER or (Strider not (short-tandem-repeat or DNA-strider* or ((water or oil or bike or bikes or biped*) adj5 strider*)))).ti. |
| 21 | 19 or 20 [ FGR & PD5-inhibitors -Strider/Tadafer trial ] |
| 22 | (randomized controlled trial/ or controlled clinical trial/ or double blind procedure/ or ((random* or (controlled adj3 (study or trial)) or placebo* or double-blind*).tw,kw. or trial.ti.)) not ((review or note).pt. or systematic review/ or review.ti.) [ RCT-filter adapted from the Cochrane ] |
| 23 | ((exp animal/ or animal experiment/ or exp animal model/ or nonhuman/) not human/) or ((pig or pigs or goat or goats* or sheep or lamb or lambs or ovine or rodent* or rabbit* or mice or mouse or murine* or rat or rats).ti. not human*.ti,ot.) [ animal filter] |
| 24 | 22 not 23 [ human RCT filter, adapted from the Cochrane ] |
| 25 | 21 and 24 [ human RCTs on FGR & PD5-inhibitors ] |
| 26 | remove duplicates from 25 [ human RCTs on FGR & PD5-inhibitors, deduplicated ] |

<u>CENTRAL</u>
Source: Cochrane Library

- Cochrane Database of Systematic Reviews. Issue 9 of 12, September 2024
- Cochrane Central Register of Controlled Trials. Issue 8 of 12, August 2024

| # | Search |
|---|---|
| 1 | ((fetal or foetal or fetus* or foetus* or intrauterin* or uterine or utero or antenat* or prenat* or early-onset or antenat* or ante-nat* or prenat* or pre-nat* or first-trimester or 1st-trimester) NEAR/6 (growth NEAR/3 (retard* or restrict*))):ti,ab,kw |
| 2 | (FGR* or IUGR*):ti,ab,kw |
| 3 | ((early or pregnancy or gestational) NEAR/2 (growth-retard* or growth-restrict*)):ti,ab,kw |

| | |
|---|---|
| 4 | ((estimat* NEAR/4 (fetal or foetal or fetus* or foetus*) NEAR/2 weight*) or EFW):ti,ab,kw |
| 5 | (((absent or revers*) NEAR/6 (enddiastol or diastol* or doppler) NEAR/2 (velocity or flow)) or AEDF* or A-EDF* or AREDF* or A-REDF*):ti,ab,kw |
| 6 | (abdom* NEAR/3 circumfer*):ti,ab,kw |
| 7 | (placent* NEAR/3 (insufficien* or dysfunct* or disorder*)):ti,ab,kw |
| 8 | (((gestational or maternal or pregnancy) NEXT (hypertension or high-blood-pressur*)) or pregnancy-induced-hypertension or (pregnancy NEAR/3 hypertensive-disorder*)):ti,ab,kw |
| 9 | (HELPP or preeclam* or eclam*):ti,ab,kw |
| 10 | 35-#9 |
| 11 | (sildenafil* or revatio or homosildenafil or hydroxyhomosildenafil or Viagra or acetildenafil or desmethylsildenafil or NCX-911 or NCX911 or UK-92480* or UK92480* or tadalafil* or cialis or ic-351 or ic351):ti,ab,kw |
| 12 | (((((phosphodiesteras* or phospho-diesteras* or PDE) NEAR/3 ("5" or V)) or phosphodiesterase5 or PDE5 or phosphodiesteraseV or PDEV) NEAR/5 (inhib* or block* or antagonist* or anti)) or PD5i* or (PD5 NEXT I) or PDVi* or PDV-I):ti,ab,kw |
| 13 | #11 or #12 |
| 14 | #10 and #13 |
| 15 | #10 and #13 in Trials |
| 16 | #15 not review:pt |

<u>WoS</u>
Source: Web of Science Core Collection

| # | Search |
|---|---|
| 1 | TS=((fetal or foetal or fetus* or foetus* or intrauterin* or uterine or utero or antenat* or prenat* or early-onset or antenat* or ante-nat* or prenat* or pre-nat* or first-trimester or 1st-trimester) NEAR/6 (growth NEAR/3 (retard* or restrict*))) |
| 2 | TS=(FGR* or IUGR*) |
| 3 | TS=((early or pregnancy or gestational) NEAR/2 (growth-retard* or growth-restrict*)) |
| 4 | TS=((estimat* NEAR/4 (fetal or foetal or fetus* or foetus*) NEAR/2 weight*) or EFW) |
| 5 | TS=(((absent or revers*) NEAR/6 (enddiastol or diastol* or doppler) NEAR/2 (velocity or flow)) or AEDF* or A-EDF* or AREDF* or A-REDF*) |
| 6 | TS=(abdom* NEAR/3 circumfer*) |
| 7 | TS=(placent* NEAR/3 (insufficien* or dysfunct* or disorder*)) |
| 8 | TS=(gestational-hypertension or maternal-hypertension or pregnancy-hypertension or pregnancy-induced-hypertension or (pregnancy NEAR/3 hypertensive-disorder*)) |
| 9 | TS=(HELPP or preeclam* or eclam*) |
| 10 | #1 or #2 or #3 or #4 or #5 or #6 or #7 or #8 or #9 |
| 11 | TS=(sildenafil* or revatio or homosildenafil or hydroxyhomosildenafil or Viagra or acetildenafil or desmethylsildenafil or NCX-911 or NCX911 or UK-92480* or UK92480* or tadalafil* or cialis or ic-351 or ic351) |
| 12 | TS=(((((phosphodiesteras* or phospho-diesteras* or PDE) NEAR/3 ("5" or V)) or phosphodiesterase5 or PDE5 or phosphodiesteraseV or PDEV) NEAR/5 (inhib* or block* or antagonist* or anti)) or PD5i* or (PD5-I) or PDVi* or PDV-I) |
| 13 | #11 or #12 |
| 14 | #10 and #13 |
| 15 | (TS=(random* or (controlled near/3 (study or trial)) or placebo* or double-blind*) or TI=trial) not TI=review |
| 16 | #14 and #15 |
| 17 | TI=(animal* or pig or pigs or goat or goats* or sheep or lamb or lambs or ovine or rodent* or rabbit* or mice or mouse or murine* or rat or rats) not TI=human* |
| 18 | #16 not #17 |

CTGOV
Source: ClinicalTrials.gov

| # | Search |
|---|---|
| | ((phosphodiesterase-5-inhibitors OR PD5i* OR PD5-i* OR PDVi* OR PDV-i* OR sildenafil OR tadalafil) AND (fetal-growth-retardation OR FGR OR HELPP OR eclampsia OR preeclampsia OR maternal-hypertension OR pregnancy-hypertension)) |

ICTRP
Source: International Clinical Trials Registry Platform (ICTRP)
Note: synonyms searched when truncation (*) isn't used

| # | Search |
|---|---|
| | Sildenafil AND fetal-growth-retardation OR sildenafil AND FGR OR sildenafil AND intrauterine-growth-retardation OR sildenafil AND IUGR OR sildenafil AND eclampsia OR sildenafil AND preeclampsia OR sildenafil AND maternal-hypertension OR tadalafil AND fetal-growth-retardation OR tadalafil AND FGR OR tadalafil AND intrauterine-growth-retardation OR tadalafil AND IUGR OR tadalafil AND eclampsia OR tadalafil AND preeclampsia OR tadalafil AND maternal-hypertension OR phosphodiesterase-5-inhibitors AND fetal-growth-retardation OR phosphodiesterase-5-inhibitors AND FGR OR phosphodiesterase-5-inhibitors AND intrauterine-growth-retardation OR phosphodiesterase-5-inhibitors AND IUGR OR phosphodiesterase-5-inhibitors AND eclampsia OR phosphodiesterase-5-inhibitors AND preeclampsia OR phosphodiesterase-5-inhibitors AND maternal-hypertension |
