## Supplementary Appendix C for "Investigating the quality, trustworthiness and integrity of published randomized trials on phosphodiesterase type 5 inhibitors for the treatment of fetal growth restriction and their impact on study findings: protocol for a systematic review with aggregate data meta-analysis and data integrity asses"

**Supplementary Appendix C.1 - Letter to author with results integrity assessments**

Subject: Concerns regarding data integrity

Dear [name corresponding author],

Our group is writing to express our concern about the following article published in the *[journal].*

[citation of article]

Previously, you were invited to contribute data to an individual participant data meta-analysis investigating the effects of phosphodiesterase type-5 inhibitors in the treatment of fetal growth restriction. Unfortunately, we have not received a response from you.

Eligible trials for this meta-analysis have been thoroughly reviewed, including an assessment of data integrity. During this process, we identified several concerns about the data integrity of the abovementioned manuscript.

[results of integrity assessments]

These findings will be presented in a manuscript that we intend to submit to *NEJM Evidence*. We kindly request your response to our findings <u>within three weeks</u>. If you require additional time, please let us know.

Should we not receive a response, or if the response is deemed inadequate, we will refer the matter to the editor of the [journal of publication] and request an investigation in accordance with COPE (Committee on Publication Ethics) guidelines[1].

Best regards, on behalf of the study team

Dr. Wessel Ganzevoort, Amsterdam University Medical Centers, Amsterdam, the Netherlands

[1]COPE Council. COPE Guidelines: Expressions of concern — English.
DOI: https://doi.org/10.24318/mw2J3661

**Supplementary Appendix C.2- Letter to editor with results integrity assessments**

Subject: Concerns regarding data integrity

Dear [name corresponding author],

Our group is writing to express our concern about the following article published in the *[journal]*.

[citation of article]

This manuscript was identified in our systematic review in the process of an IPD meta-analysis. The authors did not respond to requests for data sharing. We have scrutinized all publications and for this publication several concerns about the data integrity arose:

[results of integrity assessments]

We ask you to investigate. Given the abundance of flaws and lack of trustworthiness we ask you to retract the publication unless clear and satisfactory responses from the authors will be received. We would like to remind you that COPE guidelines[1] require an immediate expression of concern if you expect that the investigation will take more than a couple of months. We would appreciate acknowledgement of receipt of this message and follow-up messages from your side about your actions.

Best regards, on behalf of the study team

Dr. Wessel Ganzevoort, Amsterdam University Medical Centers, Amsterdam, the Netherlands

[1]COPE Council. COPE Guidelines: Expressions of concern — English.
DOI: https://doi.org/10.24318/mw2J3661
